# DIFFERENT PERSPECTIVES OF SPANISH PATIENTS AND PROFESSIONALS ON HOW A DIALYSIS UNIT SHOULD BE DESIGNED

**DOI:** 10.1101/2022.11.24.22282702

**Authors:** Maria Dolores Arenas Jiménez, Paula Manso, Fabiola Dapena, David Hernán, Jesús Portillo, Ana Balseiro, Mónica Sánchez, Raul López, Concepción Pereira, Daniel Gallego, Juan Carlos Julián, Manuel Arellano, Antonio Tombas, Iluminada Martin- Crespo, Cristina Sanz, renal foundation’s Iñigo Álvarez de Toledo work team

## Abstract

People with kidney disease on hemodialysis spend 4 hours of their lives three times a week in hemodialysis units. Although the new concept of 21st century medicine gives special prominence to the opinion of patients and family members, the reality is that this is rarely considered when establishing the requirements that a dialysis center should meet.

**Objective:** To know and compare the opinion and preferences of patients, family members and professionals regarding the design of a dialysis unit and the potential activities they believe should be carried out during the session in order to provide architects with real information for the construction of a dialysis center.

**Methods:** Anonymous and voluntary survey in electronic format addressed to patients, relatives and professionals belonging to the 18 hemodialysis centers of the renal foundation and to ALCER and its different delegations, in relation to leisure activities to be carried out in the dialysis center and preferred design of the treatment room. The results obtained between the patient-family group and the professionals were compared.

**Results:** We received 331 responses, of which 215 were from patients and family members (65%) and 116 (35%) from professionals. The most represented category among professionals was nursing (53%), followed by assistants (24%) and physicians (12.9%). A higher proportion of patients (66%) preferred rooms in groups of 10-12 patients as opposed to professionals who preferred open-plan rooms (p<0.001). The options that showed the most differences between patients and professionals were chatting with colleagues and intimacy (options most voted by patients/families), versus performing group activities and visibility (professionals).

**Conclusions:** The professionals’ view of patients’ needs does not always coincide with the patients’ perception. The inclusion of the perspective of people with kidney disease continues to be a pending issue in which we must improve both patient organizations and professionals, and the opinion of professionals and patients must be included in the design of a dialysis unit and the activities to be developed in it.

## INTRODUCTION

2,500 years ago, Lao-Tse said: “Architecture is not four walls and a roof; it is the arrangement of spaces and the spirit generated within”. The architectural design of patient care spaces has an enormous importance in aspects that are fundamental for the proper performance of the activity [1], but they have to consider also the soul of the activity that takes place in it. Hemodialysis units must be designed with the aim of achieving the greatest possible mental, physical and social rehabilitation.

Historically, the definition of what a hospital and outpatient hemodialysis unit should be like has been based on the recommendations of scientific societies and health authorities [2-4], as well as on the requirements of the different state and regional regulations [5]. This definition contemplates, among other things, the rooms it should contain, their square meters and accessibility [6-13]. In addition, aspects such as worker and patient safety [14-16] and respect for the environment and sustainability [17,18] are also considered.

Although the new concept of 21st century medicine gives special importance to the opinion of patients and family members [19], the reality is that this is rarely considered when establishing the requirements that a dialysis center must meet. People with kidney disease on hemodialysis spend 4 hours of their lives three times a week in hemodialysis units. It seems important to know their opinion about how the hemodialysis center should be designed.

The aim of this study is to know and compare the opinion and preferences of patients, family members and professionals regarding the design of a dialysis unit and the potential activities they believe should be carried out during the session in order to provide architects with real information for the construction of a dialysis center.

## METHODS

### Study design

Comparison of the results of the same questionnaire administered between November 30, 2021 and December 31, 2021 to two independent samples of the population in Spain: people with kidney disease and family members versus professionals working in the field of hemodialysis. The questionnaire asks about aspects of the design of the hemodialysis unit and activities to be developed during the hemodialysis session.

### Survey administration methodology

The survey was sent by e-mail and was addressed to relatives and patients with kidney disease affiliated to the National Federation of Associations for the Fight against Kidney Disease (ALCER) and its different delegations, as well as to patients and professionals belonging to the 18 hemodialysis centers of the Iñigo Alvarez de Toledo Renal Foundation in Spain who wished to participate. The survey was anonymous and voluntary.

### Content of the survey

The questionnaire consisted of multiple-choice and open-ended questions, which evaluated different aspects of the unit in general and some facilities such as the waiting room, dialysis room, dressing rooms, dialysis station, among others; as well as entertainment options during the session. The contents of this survey were suggested by a sample of renal patients belonging to the association of renal patients and health professionals who validated the relevance of the questions and are shown in Table 1.

**Table 1.** Questions asked and possible answers.

| Questions | Possible answers |
| --- | --- |
| How old are you? Optional answer | <25 years<br>26-50 years<br>51-65 years<br>66-75 years<br>76-80 years<br>>80 years |
| What is your gender? Optional answer | Man<br>Woman<br>Don't know/no answer |
| Categories. What is your choice? Optional answer | Physician<br>Nurse<br>Nursing assistant<br>Patients<br>Family members; relatives, caregivers<br>Patient support group (nutritionists, psychologists, social workers, sports trainers)<br>Don't know/no answer |
| What would you prefer the layout of the dialysis room stations to be? Optional answer | In an open room with all stations facing the center of the room.<br>In separate rooms for 10-12 patients<br>Single room distributed in groups of 10-12 patients<br>Don't know/no answer |
| What do you value most about the dialysis room? Multiple-choice and open-ended answers | To allow talking with other dialysis patients during the session.<br>That allows for group activities<br>That allows isolation from others and privacy<br>Good visibility from the nursing control |
| What entertainment/activity options do you find best during hemodialysis? Multiple choice and open-ended answers | Movies/documentaries/series<br>Music<br>Group discussions<br>Board games/bingo<br>Virtual reality<br>Individual activities (Painting, drawing, crafts.)<br>Reading<br>Educational workshops<br>Sports |
| How do you like the colors of the dialysis unit? Multiple choice and open-ended answers | Warm colors<br>Pastel colors<br>White<br>Bright colors<br>Green/blue |
| How would you rate the presence of natural plants in the unit? Numerical scale from 1 to 5 | Not very important<br>Somewhat important<br>Regularly important<br>Fairly important<br>Very important<br>Don't know/no answer |
| How important is privacy to you at certain times during dialysis (e.g., catheter connection, dizziness, sleeping...) Numerical scale 1 to 5 | Not very important<br>Somewhat important<br>Regularly important<br>Fairly important<br>Very important<br>Don't know/no answer |
| What would you most value your dialysis station having? Multiple choice and open-ended answers | Table<br>Socket for your use<br>Shelf for personal belongings<br>Individual light<br>Bag hanger<br>Don't know/no answer |
| The entrance to the unit is a very important place How can we make a waiting room more comfortable? Open answer | Open answer |
| What would you like the locker room to look like? Open answer | Open answer |
| Where would you like the nursing control to be located? Optional answer | Far from the dialysis station, but with visibility<br>Near the dialysis station<br>Don't know / no answer |
| What would you change in the dialysis center you know? | Open answer |
| What would you highlight as the best thing about the dialysis center that you know? | Open answer |
| What would your ideal dialysis center look like? | Open answer |
| As a professional, what would make your day-to-day life at the center easier and safe? | Open answer |

### Variables

In addition to the specific questionnaire, some minimum demographic variables were collected: age, sex, person completing the survey: patient, family member or professional, and different professional categories: physician, nurse, nursing assistant, member of the renal patient support group (nutritionists, social workers, psychologists, sports technicians).

### Statistical analysis

Numerical variables were described with the mean and standard deviation, while categorical variables were described with the number and percentage of subjects. Comparisons between groups were performed with Student’s t-test for continuous variables and the chi-square test for categorical variables. In all tests, the significance level was considered to be p<0.05. All the analyses were performed using the SPSS 28 statistical program for Window

### Ethical considerations

This analysis followed the regulations of the European Union law on data protection and privacy for all persons within the European Union (GDPR / 2018), the Declaration of Helsinki on Ethical Principles for Medical Research Involving Human Subjects. Before answering the survey, participants were given access to the following information sheet. Answering the survey and sending it was considered as the participant’s acceptance to participate in the study. No personal data were collected and there was no possibility of knowing the identity of the participants.

## RESULTS

A total of 331 answers were received, of which 215 were from patients and family members (65%) and 116 (35%) from professionals. Table 2 shows the description of patients and professionals. The two populations were demographically different: older patients and predominantly male (60%) and younger professionals with almost 80% women. The most represented category among professionals was nursing (53%), followed by assistants (24%) and physicians (12.9%).

**Table 2.** Characteristics of the study populations.

|  |  | Patients/relatives<br>N= 215 | Professionals<br>N=116 | Total<br>N=331 | p |
| --- | --- | --- | --- | --- | --- |
| How old are you?<br>Optional answer | <25 years | 4(1,9%) | 7 (6%) | 11 (3,3%) | <0,001 |
|  | 26-50 years | 78 (36,3%) | 91 (78,4%) | 169(51,1%) |  |
|  | 51-65 years | 88(40,9%) | 17(14,7%) | 105(31,7%) |  |
|  | 66-75 years | 34 (15,8%) | 1 (0,9%) | 35 (10,6%) |  |
|  | 76-80 years | 7 (3,3%) | 0 ( 0%) | 7 ( 2,1%) |  |
|  | >80 years | 4(1,9%) | 0 (0%) | 4(1,2%) |  |
| What is your gender?<br>Optional answer | Man | 129 (60,3%) | 24(20,7%) | 153(46,4%) | <0,001 |
|  | Woman | 85 (39,7%) | 92(79,3%) | 177(53,6%) |  |
| Categories. What is your<br>choice? Optional answer | Physician | 0(0%) | 15(12,9%) | 15(4,5%) | <0,001 |
|  | Nurse | 0 (0%) | 62(53,4%) | 62(18,7%) |  |
|  | Nursing assistant | 0 (0%) | 28(24,1%) | 28(8,5%) |  |
|  | Patients | 202(94%) | 0(0%) | 202(61%) |  |
|  | Family members;<br>relatives, caregivers | 13(6%) | 0(0%) | 13(3,9%) |  |
|  | Patient support group | 0(0%) | 5(4,3%) | 5(1,5%) |  |
|  | Don't know/no answer | 0(0%) | 6(5,2%) | 6(1,8%) |  |

Patient support group: nutritionists, psychologists, social workers, sports trainers

### Comparison between patients and professionals

Table 3 shows the comparison between patients/ relatives and professionals in relation to hemodialysis room design preferences and the activities to be carried out during the dialysis session. Two thirds of the patients preferred small rooms with 10-12 places, while only one third of the professionals opted for this alternative (p<0.001). The options that showed the greatest differences between patients and professionals were chatting with colleagues and intimacy (options most voted by patients/families), as opposed to group activities and visibility (professionals).

**Table 3.** Preferences of patients and professionals in the design of the hemodialysis room and activities to be carried out during dialysis.

|  | Patients/relatives<br>N= 215 | Professionals<br>N=116 | Total<br>N=331 | p |
| --- | --- | --- | --- | --- |
| What would you prefer the layout of the dialysis room stations to be? Optional answer |  |  |  |  |
| In an open room with all stations facing the center of the room. | 76 (35,3%) | 70(60,3%) | 146(44,1%) | <0,001 |
| In groups of 10-12 patients | 130(60,4%) | 44(37,9%) | 174(52,5%) |  |
| a) Single room distributed in groups of 10-12 patients | 48(22,3%) | 21 (18,1%) | 69 20,8%) |  |
| b) In separate rooms for 10-12 patients | 82(38,1%) | 23(19,8%) | 105 (31,7%) |  |
| Don't know/no answer | 9(4,1%) | 2(1,7%) | 11(3,3%) |  |
| What do you value most about the dialysis room?<br>Multiple-choice and open-ended answers |  |  |  |  |
| To allow talking with other dialysis patients during the session. | 112 (52%) | 45(38,7%) | 157(47,4%) | <0,001 |
| That allows for group activities | 36(16,7%) | 47 (40,5%) | 83(25%) |  |
| That allows isolation from others and privacy | 67(31,1%) | 9(7,7%) | 76(22,9%) |  |
| Good visibility from the nursing control | 0(0%) | 15(12,9%) | 15(4,5%) |  |
| ¿ What entertainment/activity options do you find best during hemodialysis? Multiple choice and open-ended answers |  |  |  |  |
| Movies/documentaries/series | 155/217(71,4%) | 62/217(28,5%) | 217/331 (65,5%) | <0,001 |
| Music | 81/147 (37,7%) | 66/147 (56,9%) | 147/331(44,4%) | <0,001 |
| Group discussions | 51/95(23,7%) | 44/95(37,9%) | 95/331(28,7%) | 0,006 |
| Board games/bingo | 36/88 (16,7%) | 52/88(44,8%) | 88/331(26,5%) | <0,001 |
| Virtual reality | 25/42(11,6%) | 17/42(14,7%) | 42/331(12,6%) | 0,266 |
| Individual activities (Painting, drawing, crafts.) | 19/41 (8,8%) | 22/41(19%) | 41/331(12,3%) | 0,007 |
| Reading | 11/8(5,1%) | 5/16(4,3%) | 16/331(4,8%) | 0,487 |
| Educational workshops | 2/8 (0,9%) | 6/8(5,2%) | 8/331(2,4%) | 0,024 |
| How do you like the colors of the dialysis unit? Multiple choice and open-ended answers |  |  |  |  |
| Warm colors | 39 (18,1%) | 26(22,4%) | 65(19,6%) | 0,350 |
| Pastel colors | 46 (21,4%) | 13(11,2%) | 59(17,8%) | 0,021 |
| White | 116 (54%) | 61 (52,6%) | 177(53,5%) | 0,812 |
| Bright colors | 53(24,7%) | 36(31%) | 89(26,9%) | 0,211 |
| Green/blue | 15(7%) | 12(10,3%) | 27(8,2%) | 0,285 |
| How would you rate the presence of natural plants in the unit? Numerical scale from 1 to 5 |  |  |  |  |
| Not very important | 10 (4,7%) | 12 (10,3%) | 22(6,6%) | <0,001 |
| Somewhat important | 10 (4,7%) | 6 (5,2%) | 16(4,8%) |  |
| Regularly important | 38 (17,7%) | 24 (20,7%) | 62(18,7%) |  |
| Fairly important | 30 (14 %) | 32 (27,6%) | 62(18,7%) |  |
| Very important | 122(56,7%) | 40 (34,5%) | 162(48,9%) |  |
| Don't know/no answer | 5 (2,3%) | 2(1,7%) | 7(2,1%) |  |
| How important is it for you to have privacy at certain times during dialysis (e.g. catheter connection, dizziness, sleep, etc.)? |  |  |  |  |
| Not very important | 24 (11,2%) | 2 (1,7%) | 26(7,9%) | <0,001 |
| Somewhat important | 16 (7,4%) | 1(0,9%) | 17(5,1%) |  |
| Regularly important | 28 (13%) | 10 (8,6%) | 38(11,5%) |  |
| Fairly important | 26 (12,1 %) | 31 (26,7%) | 57(17,2%) |  |
| Very important | 117(54,4%) | 69 (59,5%) | 162(56,2%) |  |
| Don't know/no answer | 4(1,9%) | 3(2,6%) | 7(2,1%) |  |
| What would you most value your dialysis station having? Multiple choice and open-ended answers |  |  |  |  |
| Table | 94(43,7%) | 50(43,1%) | 144(43,5%) | 0,914 |
| Shelf for personal belongings | 78(36,3%) | 52(44,8%) | 130(39,3%) | 0,129 |
| Individual light | 70(32,6%) | 55(47,4%) | 125(37,8%) | 0,009 |
| Socket for use (cell phone, tablet, computer) | 135(62,8%) | 67(57,8%) | 202(61%) | 0,370 |
| Bag hanger | 36(16,7%) | 24(20,7%) | 60(18,1%) | 0,374 |
| Where would you like the nursing control to be located? Optional answer |  |  |  |  |
| Far from the dialysis station, but with visibility | 84 (39,1%) | 58(50%) | 142(42,9%) | <0,001 |
| Near the dialysis station | 55(25,6%) | 40(34,5%) | 95(28,7%) |  |
| Don't know / no answer | 76 (35,3%) | 18(15,5%) | 94(28,4%) |  |

**Table 4.** Compilation of the main responses to the questions asked, ordered from most to least frequent responses.

| Questions | Open answers |
| --- | --- |
| What entertainment/activity options do you find best during hemodialysis? | "The best thing would be not to have to go to dialysis"; "Heart magazines, travel magazines..."; "that the group talks are not about our disease but about other topics"; "it depends on how I feel"; "Sleep"; "Cooking classes"; "Mindfulness". |
| What do you value most in the dialysis room? | "Sufficient space"; "More comfortable chairs"; "Natural light" ; " no air conditioning jet falling on top"; "Wifi connection"; "Individual headphones"; Individual television"; "Adequate attention from doctors and staff"; "Adequate attention from doctors and staff". |
| The entrance to the unit is a very important place How can we make a waiting room more comfortable? Open answer | "Large, bright and comfortable waiting room"; "comfortable armchairs, not chairs"; "special space for wheelchairs"; "not like a hospital waiting room"; comfortable armchairs, not chairs"; "special space for wheelchairs"; "not like a hospital waiting room"; "piped music, internet access, television, magazines, newspapers, entertainment elements". |
| What would you like the locker room to look like? Open answer | "spacious, comfortable, air-conditioned, clean and with benches for changing"; "with large, individual, private lockers with hangers"; "that allow sufficient privacy" |
| What would you change in the dialysis center you know? | **Patients** : "larger waiting room"; "more intimacy"; "air conditioning"; "no activities during the session"; "less noise in the room"; "white light, I would put warm and dim light"; "armchairs are uncomfortable, there is nowhere to leave the bag"; "shared TVs"; individual light"; "more seats in the waiting room and changing rooms"; "more interaction"; "less people in the room"; "more contact with the doctors"; "administer solids and liquids within their possibilities according to their health"; "waiting times for ambulances"; "more contact with the doctors"; "more contact with the doctors"; "administer solids and liquids within their possibilities |
|  | according to their health"; "waiting times for ambulances".<br>**Professionals:** "larger and better distributed storerooms and changing rooms"; "more space between patients"; "medical office close to the ward"; "more staff";<br>"malfunctioning of ambulances and waiting times"; "layout and colors"; "automation of information systems with data dump". |
| What would you highlight as the best thing about the dialysis center that you know? | **Patients:** "The personal quality and the care provided by the professionals".<br>**Professionals:** "The team of professionals and the treatment"; "the spaciousness"; "the natural light". |
| What would your ideal dialysis center look like? | **Patients:** "spacious, at street level, warm, offering privacy in various sectors (offices, rooms, dressing rooms); "that makes you feel comfortable and safe"; "that in case of an emergency in the room has some kind of rail with curtain and cover so that we do not see the other patients, that leaves us bad body"; "close to home and with activities to be entertained"; natural light and possibility of dim light after the connection"; "a center that looks less like a hospital"; "with wifi connection"; "with very comfortable armchairs or beds"; "with views"; "with color, plants, internet, music"; "with good professionals"; "without waiting"; "with tablets and computers"; "with good professionals"; "without waiting"; "with tablets and computers"; "with a good quality of life"; "with a good quality of life"; "with a good quality of life"; "with a good quality of life"; "with good quality of life".<br>**Professionals:** "Accessible, spacious, with lots of natural light, good equipment and staff"; "space for wheelchairs"; "good air conditioning"; "with good team atmosphere"; "with a good team atmosphere"; "with a good atmosphere"; "with a good team atmosphere"; with good team atmosphere"; "bright at connection and dim light afterwards"; "latest technology in monitors and security". |
| As a professional, what would make your day-to-day life at the center easier and safe? | "Crane to move patients and adapted scales"; "Well-differentiated circuits"; "Centralized bathroom to avoid carrying bottles"; "Spaciousness in the room, changing rooms and offices"; "Larger changing rooms"; "Space between armchairs"; "Proximity to public transport and parking"; "Fewer patients per nurse, more staff"; "Continuing education"; "Intimate space to talk to patients"; "Identification of staff and explanatory leaflets for patients"; "Computers, tablets, wifi". "Differentiated circuits: ideally it should have two entrances to be able to separate the accesses of people from those of supplies, maintenance or waste"; "hallway and circulation areas must be wide with as few turns as possible, and if these exist, they must allow easy passage of a stretcher or bed (at least 2 meters)"; "wheelchair parking"; supervisory, medical and office/break spaces have direct visual and physical contact with the treatment room, as well as easy access to it" |

### Influence of age or gender in patients

Age over 65, but not gender, influenced some of the activities chosen by patients (76% of those over 65 choose to watch movies and documentaries vs. 57.8% of those under 65 (p=0.015). and 26.5% of those over 65 choose to chat with friends vs. 13.3% of those under 65, (p= 0.045) and virtual reality was chosen by 14% of those over 65 and only 2.2% of those under 65 years of age (p=0.016)

## DISCUSSION

The center guidelines that establish how the architectural design of hemodialysis units should be in Spain [4], do not reflect the opinions and preferences of people with kidney disease on dialysis about what the ideal dialysis center should be like. People with kidney disease are the focus of hemodialysis units [20,21] and these should be designed to improve their experience [22,23], so their opinions should be considered when giving indications on how to design them. This is the first survey in Spain that attempts to approach both the opinions of the patients and the professionals working in the units. Many of the answers obtained in the survey coincide with the recommendations made in the Spanish guidelines for hemodialysis centers [4], such as the need for the center to provide comfort, privacy, spaciousness, natural light, ventilation, and for a domestic environment to predominate over a hospital environment, as evidenced by the phrase “a center that looks less like a hospital”, which increases the validity of these guidelines [4].

One of the novelties of this survey is that it gives a voice to the people who dialyze and work in dialysis units in more specific aspects of structure and operation such as the layout of the dialysis room, the color of the walls, the presence of natural plants, the location of the nursing control in relation to the dialysis station, the entertainment options during the session, types of lighting and elements that they consider necessary to be in their dialysis station such as sockets, table, bag hanger, etc. and attempts to translate into practice the broad and ill-defined terms we commonly use, such as comfort or privacy.

Although aspects such as air conditioning, natural light and a quiet environment with absence of noise [24] were equally valued by patients and professionals, there are other characteristics in which the opinions and/or preferences of professionals do not coincide with those of people with kidney disease, such as the layout of the dialysis room or the activities to be performed during the session.

Most patients prefer smaller rooms or rooms distributed in groups of 10-12 patients, which allow them a better relationship with their colleagues and greater intimacy, while a greater number of professionals prefer large, diaphanous rooms. Probably all opinions, although different, should be considered when designing a center with smaller patient modules, but respecting the necessary amplitude required by the professionals. In general, patients attach greater importance to comfort, the relationship in the room with other colleagues and intimacy, while professionals prioritize patient control and safety.

The different responses found in patients and professionals further support, if possible, the need to include the latter, as well as their relatives and caregivers, in many processes that we take for granted and to listen to their views and preferences to achieve the goal of improving the patient experience.

Probably all opinions, although different, should be considered when designing and building a center, with smaller patient modules, but respecting the necessary amplitude required by the professionals.

Early involvement of physicians and nurses in the construction of health care facilities [25,26] is not new, and has been shown to reduce initial and operating costs, improve facility performance, and provide a safe environment with the best solutions [25,26]. This survey allowed the architects to modify the initial design of the unit and adapt it to the preferences of patients and professionals. The single, open-plan room initially proposed was converted into separate 12-place rooms with movable panels. Interior gardens were placed to allow the inclusion of plants as a decorative element and the white lights were changed to warm ones.

Especially relevant is the fact that, although the questions are directed to the interior design of the dialysis center in architectural terms, both professionals and patients, when asked open-ended questions: “What would you highlight as the best thing about the dialysis center you know? What would your ideal center be like?”, mention the human quality, the care provided by professionals, and the work team and work environment, as well as highlighting aspects that have nothing to do with the architecture itself, such as waiting times or staff training and ratios. Our analysis, like other studies [27,28], highlights the need to humanize infrastructures, since they are a vehicle for humanizing care, designing spaces where our patients feel comfortable while receiving their treatment.

On the other hand, this survey also shows the importance of new technologies (Tablet, Wifi, security, computers, data transfer, etc.) in improving the experience of both patients and workers, which we must necessarily incorporate in the design of new units and adapt existing ones. It is true that one of the limitations of this study is the fact that the sample may not be representative of the entire dialysis population, since it is likely that people who do not have access to e-mail are not represented, but at least they are in the population that answered the survey.

Some open responses, both from professionals and patients, refer to “ambulance waiting times”. Transportation and waiting times in dialysis units have been shown to have an influence, not only on the patient experience, but also on the higher mortality and lower quality of life associated with longer waiting and travel times [29].

In conclusion, the vision of professionals about the needs of patients does not always coincide with their perception, so it is necessary to ask people with kidney disease directly and listen actively, so that they can reflect their preferences and unmet needs. The inclusion of the perspective of people with kidney disease continues to be a pending issue in which we must improve. The opinion of professionals and patients must be included in the design of a dialysis unit and the activities to be carried out in it to improve the experience in it.

## Data Availability

All relevant data are within the manuscript and its Supporting Information files.

## Financing

This manuscript has not received any funding.

## APPENDIX Renal foundation’s Iñigo Álvarez de Toledo work team

Isabel Gonzalez

Marina Burgos

Maria Luz Sánchez

Marta SanJuan

Luis Nieto

Ramiro Cazar

Jose Guerrero -

Adriana Iglesias

Lola Piña

Maria Lopez Picasso

Monica Pereira;

Maria delgado

Solmar rodriguez

